# Simplifying Daily Cortisol Cycle Analysis: Validation and Benchmarking of the Cortisol Sine Score Against Cosinor and JTK_CYCLE models

**DOI:** 10.64898/2026.02.23.26346831

**Authors:** Simone Anzà, Bruce A. Rosa, Max P. Herzberg, Giljae Lee, Erik D. Herzog, Peinan Zhao, Sarah K. England, I. Malick Ndao, John Martin, Christopher D. Smyser, Cynthia E. Rogers, Deanna M. Barch, Caroline Hoyniak, Ronald McCarthy, Joan Luby, Barbara B. Warner, Makedonka Mitreva

## Abstract

The daily cortisol cycle is a critical indicator of hypothalamic-pituitary-adrenal (HPA) axis function. The current analytical approaches produce several outputs difficult to integrate into simple statistical models, clinical workflows, and ML/AI pipelines requiring single-value inputs. We developed the Cortisol Sine Score (CSS), a model-free scalar metric that quantifies daily cortisol exposure by computing a weighted sum of cortisol measurements across the day, using sine-transformed time-of-day weights. The CSS produces positive values for morning-dominant patterns, negative values for evening-shifted profiles, and near-zero values for flattened rhythms characteristic of chronic stress and circadian disruption. We validated the CSS performance in 3,006 samples from 501 pregnant women enrolled in the March of Dimes program, with cortisol values measured at 6 time points per day collected during the second trimester of pregnancy. The CSS showed strong correlations with observed and model-estimated amplitude and acrophase from Cosinor regression and JTK_CYCLE approaches, with excellent classifying performance (AUC=0.89, high versus low). The CSS successfully captured established associations between social disadvantage and cortisol dysregulation, and demonstrated utility in predicting gut microbiome composition in metagenomic analyses. Importantly, the CSS maintains excellent fidelity to the full 6-sample protocol with as few as 3-4 daily measurements. The 4-sample protocol achieves great performance (r = 0.952, MAE = 0.087) while reducing participant burden. The 06:00 time point was identified as essential for accurate CSS quantification. The CSS bridges the gap between circadian analysis and practical implementation by providing a simple, interpretable, and robust assessment of cortisol daily cycle in large-scale epidemiological studies, clinical screening, and biomedical sensors.

**Highlights:**

- Current state-of-the-art approaches estimating the daily cortisol exposures produce multi-output information difficult to implement in simple statistical analyses or ML/AI multi-omics approaches
- Cortisol Sine Score is a novel model-free scalar metric expressing cortisol daily exposure and rhythmicity (morning vs evening exposure)
- Cortisol Sine Score was validated using 3006 salivary samples from clinical data and golden standards in circadian analyses such as Cosinor and JTK_CYCLE
- Cortisol Sine Score was the top performer in our benchmarking approach predicting association with social disadvantage and gut microbiome composition
- Reliable with 3-4 daily samples, reducing participant burden
- Open-source R package CortSineScore democratizes cortisol cycle analysis

## 1. Introduction

The cortisol daily cycle follows a well-characterized pattern regulated by the circadian system: a sharp peak in the early morning, followed by a gradual decline with a nadir in the evening/early night. Alterations of this cycle have been consistently linked to chronic stress disorders, detrimental health, psychiatric disorders, metabolic dysfunction, and disrupted sleep-wake cycles[1–3].

To better understand hypothalamic-pituitary-adrenal (HPA) axis functionality via characterization of cortisol regulation, a variety of metrics are used to summarize different aspects of the cortisol daily cycle. The Area Under the Curve with respect to ground (AUCg) quantifies total cortisol output over a period of 24 hours and is often used as a measure of cumulative exposure[4]. Peak values typically occur in the early morning and are used to index the magnitude of activation of the HPA-axis. Amplitude, a measure of rhythm strength, reflects the difference between peak and trough levels, while acrophase denotes the time of peak secretion, indicating phase alignment with the circadian clock. Other approaches characterize rhythmicity by testing whether cortisol exposure follows a predictable 24-hour waveform, or test reactivity by assessing cortisol response to acute challenges or transitions[5,6]. These parameters capture different biological features of the HPA-axis, but no single metric comprehensively describes the system’s function across different dimensions.

Despite the availability of multiple metrics, capturing the overall shape and integrity of the cortisol daily cycle in a simple and single interpretable value remains an unsolved challenge. While AUC provides a single metric indexing cumulative exposure, it does not provide information about the circadian alignment of cortisol secretion. Methods such as Cosinor regression[5,7] and the Jonckheere-Terpstra-Kendall Cycle (JTK_CYCLE)[6], are powerful approaches for parameter estimation (e.g., amplitude, acrophase) or rhythmicity tests. However, these approaches produce multiple outputs that may not be easily incorporated into downstream statistical models like classic regressions, mediation analyses, or ML/AI frameworks requiring single input parameterization. The Cosinor model is a parametric regression-based method that estimates amplitude, mesor, and acrophase, fitting a sinusoidal waveform with a fixed period (typically 24 hours)[5,7]. The JTK_CYCLE approach is a non-parametric test originally developed for high-throughput time-series data like gene expression. It detects rhythmicity by comparing data to a bank of reference templates at varying phases using rank-based correlation statistics, and it calculates a JTK test statistic to evaluate monotonic or rhythmic trends. The outputs include: *P*-values for rhythmicity, estimated period and phase, and estimated amplitude [6]. Unlike Cosinor, the JTK_CYCLE does not assume a parametric form or waveform shape, making it robust to non-sinusoidal rhythmic patterns. However, it is less informative in terms of generating continuous variables for downstream modeling and is primarily used to detect whether a rhythm exists. By relying on multi-output models, the current state-of-the-art methods are difficult to implement in large epidemiological studies, clinical settings or ML/AI, requiring single numeric input parameterization. There is a critical need for a unified, scalar metric that can approximate cortisol daily cycle features such as timing, alignment, and directionality without sacrificing interpretability or applicability in statistical analysis. To address this limitation, we developed the Cortisol Sine Score (CSS): a simple, model-free, scalar metric that collapses both the amount and timing of cortisol into a single, biologically interpretable index of rhythm alignment. Here, we present the validation of our new CSS approach and comparison of its performance to standards in the field.

### 1.1 Biological Rationale

Cortisol secretion is tightly regulated by the circadian system, exhibiting a high-amplitude diurnal rhythm that typically peaks shortly after waking up and declines progressively throughout the day. This morning-dominant pattern is a hallmark of HPA-axis integrity and healthy circadian alignment [8]. Conversely, flattened, delayed, or inverted secretion profiles have been widely observed in contexts of chronic stress, psychiatric illness (e.g., depression), metabolic dysregulation, and adverse environmental exposures[3,9]. The value of the CSS lies in its interpretability and applicability: positive values reflect morning-aligned exposure, typical of a healthy cortisol daily cycle, negative values suggest delayed or inverted rhythms, while near-zero values indicate flattened or dysregulated patterns. This simplification enables the integration of complex cortisol dynamics into standard statistical pipelines without the need for multi-parameter modeling. Broadly speaking, the CSS answers the question: is cortisol secretion early-day dominant (circadian-aligned), flattened, or later-shifted and therefore potentially misaligned?

### 1.2 Mathematical Rationale

The CSS design was inspired by the Fourier decomposition principles [10], in which biological rhythms are projected onto sine wave templates. However, unlike full Fourier series or Cosinor fitting, the CSS collapses the time series into a single weighted sum, using sine-transformed time-of-day weights to distinguish between cortisol exposure during the early (positive contribution) versus late (negative contribution) times of the day. This operation is mathematically expressed as a dot product between the individual’s cortisol time series and a canonical 24-hour sine wave. Specifically, the CSS is a scalar metric designed to quantify the alignment and directionality of diurnal cortisol secretion patterns relative to the 24-hour circadian cycle. For each individual, the CSS is computed as a weighted sum of cortisol values collected at different timepoints, where weights are derived from the sine of the time of day (converted into radians on a 24-hour circle):

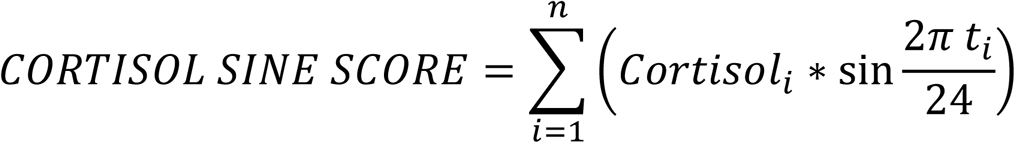

Here, *n* represents the number of timepoints (samples collected over 24 hours), *Cortisol* is the salivary cortisol concentration (e.g., ng/mL) at specific timepoint *i*, and *t_i_* is the clock time in hours. This transformation projects each cortisol value onto a 24-hour sine wave such that the early morning timepoints contribute positively to the index, the afternoon/evening timepoints contribute negatively, and positive or negative values near midday (12:00) contribute minimally. It is designed to distinguish among these patterns by leveraging the natural structure of the 24-hour cycle and weighting the cortisol values such that morning cortisol exposure (from 00:00 to 12:00) contributes positively to the CSS, and evening cortisol exposure (from 12:00 to 00:00) contributes negatively. Hence, the CSS peaks when the sine function peaks (morning-dominant), and it decreases if cortisol dips when the sine function dips (evening-dominant). If the cortisol exposure is equally distributed across the day (e.g., flattened cycle), these contributions cancel out, producing near-zero values. Importantly, the CSS is model-free, requiring no model fitting, making it especially suitable for large-scale studies, regression models, or clinical applications requiring a single interpretable outcome.

To validate this novel metric, the CSS was compared against two widely used approaches: Cosinor-regression and JTK_CYCLE (described above). We evaluated CSS performance in recapitulating rhythmic features captured by Cosinor and JTK_CYCLE including amplitude and acrophase (peak timing), and we benchmarked its ability to replicate well-known biological associations with cortisol disruption against other metrics (e.g., AUCg, MESOR, Acrophase, Amplitude), and provided an example on how to use CSS in multi-omics studies. We created the open-source R package *CortSineScore,* which performs the cortisol sine score estimation, and is publicly available for use (https://github.com/simone-anza/CortSineScore). Together, our work introduces and validates the CSS as a compact, biologically interpretable, and practical alternative for modeling cortisol daily cycle patterns in large-scale studies and applied contexts where a single scalar input is preferred or required.

## 2 Materials and Methods

### 2.1 Participants, salivary cortisol sampling and processing

This study includes data collected from 830 pregnant women from the March of Dimes cohort[11]. We included only subjects (n = 501) with cortisol levels measured at six time points within a single day: 02:00, 06:00, 10:00, 14:00, 18:00, 22:00. The participants provided salivary samples using swabs and Salivette tubes (SAR-511534500; Sarstedt) every 4 hours for 24 hours in each trimester during pregnancy; details on salivary sample collection, storage, data cleaning, preprocessing, and AUCg estimations are provided in Herzberg et al, 2023 [12]. Final validation and benchmarking of the CSS involved the use of 3006 samples from the second trimester only.

To determine the minimum number of daily cortisol measurements required to estimate the CSS reliably, we systematically evaluated all possible combinations of the six time points. We analyzed 63 combinations in total: 6 single-sample, 15 two-sample, 20 three-sample, 15 four-sample, 6 five-sample, and 1 six-sample. For each combination, we calculated the CSS using only the cortisol values and sine weights corresponding to the selected time-point samples. To ensure comparability across different sampling schemes, we applied min-max normalization to rescale all CSS values to the range [-1, 1]. Each reduced sampling CSS metric was compared to the full 6-sample CSS metric as a gold standard using the Pearson’s correlation coefficient and mean absolute error (MAE). We used the threshold of Pearson’s r > 0.9 and MAE < 0.1 corresponding to < 5% error. After we identified the minimum number of daily cortisol measurements and built the corresponding CSS metric, we benchmarked its performance versus the 6-timepoint CSS metric.

### 2.2 Cortisol Sine Score validation

We validated the CSS by testing for correlations between CSS and other metrics derived by state-of-the-art approaches used in circadian rhythm and cyclicity analyses such as the (i) Cosinor regression modelling, and (ii) the JTK_CYCLE statistical approach. Cosinor models used in R (e.g., via *cosinor* or *cosinor2* packages) fit a parametric sinusoidal curve to time-series data, typically assuming a single-component cosine wave to model circadian rhythms [5]. The standard cosinor model is:

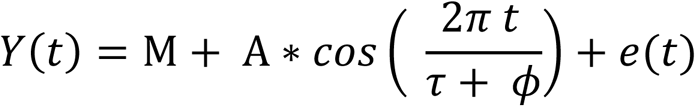

where M represents a rhythm-adjusted mean known as the Midline Statistic Of Rhythm (MESOR), A represents a measure of half the extent of predictable variation within a cycle (amplitude), ϕ represents the time of overall high values recurring in each cycle (acrophase), τ is cycle duration (period), and e(t) represents the error term [5]. The cosinor regression was applied using the *cosinor* R package [7]. For each participant, a regression model was fit under the following assumptions [5,7]:

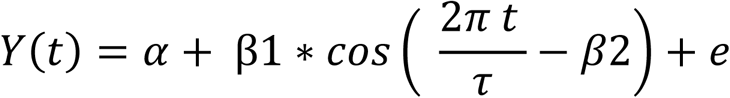

Where α represents the MESOR, β1 is the amplitude, β2 is the acrophase (peak time) and ε represents the residual error. This model is nonlinear and it’s transformed into a linear model using trigonometric identities [5,7]. In contrast, the JTK_CYCLE algorithm is primarily used to detect whether a rhythm exists. It calculates a JTK_CYCLE test statistic to evaluate monotonic or rhythmic trends, and outputs include: P-values for rhythmicity (unadjusted and FDR-corrected), estimated period and phase lag, and estimated amplitude [6]. The R packages *cosinor* and *MetaCycle* were used to estimate amplitude and acrophase according to the cosinor and JTK_CYCLE approaches, respectively.

To validate the CSS approach, we fitted statistical models on CSS versus observed and estimated amplitude and acrophase via cosinor, and JTK_CYCLE estimation. Specifically, we tested for association between CSS and: i) observed amplitude, ii) cosinor-estimated amplitude, and iii) JTK_CYCLE-estimated amplitude by fitting 3 different linear mixed models using the R package *glmmTMB* [13]. Additionally, we implemented Receiver Operating Characteristic (ROC) curve analyses and evaluated the ability of the CSS treated as a continuous predictor to accurately classify individuals with high (top 25% quartile) versus low (bottom 25% quartile) amplitude by plotting the true positive rate (recall) against the false positive rate (precision). The resulting curve provides a visual and quantitative measure of how well a given metric can distinguish between binary classes. We summarized the performance using the area under the ROC curve (AUC). We computed ROC curves and AUCs using the R package *pROC* [14].

Because acrophase is a temporal variable and has a circular nature, we performed two different statistical approaches to test for correlation between CSS and acrophase: a) categorical linear mixed models, and b) generalized additive models. First, we divided our population into tertiles according to CSS values, and then we ran linear mixed models to test for CSS-categories differences (high, mid, low) in observed, cosinor-estimated, and JTK_CYCLE-estimated peak hour time (acrophase). Second, we fitted generalized additive models (GAMs) with a cyclic smoothing spline; this approach allows for flexible modelling of non-linear associations while accounting for the circular nature of time-of-day data and captures how CSS varies as a function of peak timing, potentially reflecting phase alignment or misalignment with the expected diurnal cortisol rhythm. Separate GAMs were fitted using (i) observed cortisol Acrophase, (ii) Cosinor-estimated Acrophase, and (iii) JTK_CYCLE-estimated Acrophase as predictors of CSS. We fitted each GAM with a cyclic cubic spline basis with 6 basis functions (k = 6) and a period of 24 hours to reflect circadian periodicity using the package *mgcv* [15,16]. Smoothing parameters were estimated using restricted maximum likelihood (REML).

### 2.3 Cortisol Sine Score benchmark

We benchmarked the CSS performance via across-method comparison of Spearman correlations between different cortisol daily cycle metrics and Social Disadvantage (SD). The SD variable integrates multiple indicators across domains of socioeconomic status, neighborhood context, and healthcare access and includes factors such as Income-to-needs ratio, National Area Deprivation Index percentile, Healthy Eating Index, Maternal educational attainment, and Health insurance status[17]. The SD has been previously established to have associations with the daily cortisol cycle[3,12,18–20] . Our benchmarking framework included the following variables: AUCg, MESOR, observed amplitude, observed acrophase, Cosinor-estimated amplitude, JTK_CYCLE-estimated amplitude, atypicality, amplitude atypicality, and acrophase atypicality.

We also created the *Atypicality* metric (composed of amplitude atypicality and acrophase atypicality) as another simple scalar metric used to compare the results of CSS with other measures with similar characteristics (e.g., simple scalar metrics of combined information). In this analysis, *Atypicality* measures how much an individual’s cortisol daily cycle deviates from the group-average cycle. We used two rhythm parameters: amplitude and acrophase. A daily cycle is considered more atypical if its amplitude and/or acrophase are far from the group-average values of amplitude and acrophase. The Combined Atypicality is a single score summarizing both deviations. First, we computed the group averages: mean amplitude and mean acrophase. While the mean amplitude is the mean of all individuals’ amplitudes, the mean acrophase is the circular mean (time is cyclical) of all individuals’ peak times. Therefore, we converted each observed individual acrophases into radians θ and then estimated the average cosine (C) and sine (S) components:

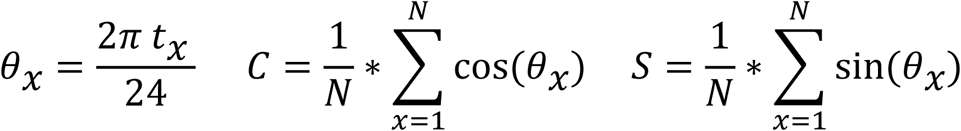

where *t_x_* is the acrophase in hours for individual *x*; and then estimated the mean angle and the mean acrophase in hours as follows:

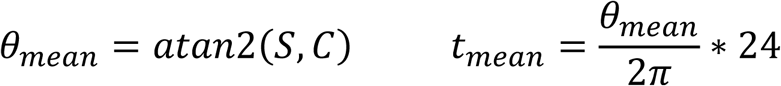

We calculated amplitude and acrophase atypicality as follows:

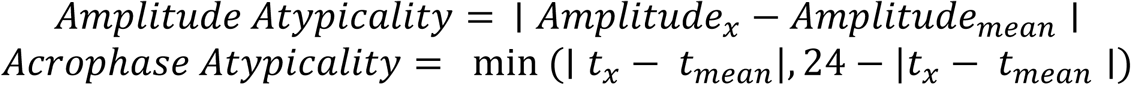

with acrophase atypicality as the circular distance (shortest distance around the 24-hour clock) from the mean acrophase, where *t_x_* is the individual acrophase, and *t_mean_* is the group-average acrophase. Since amplitude and acrophase are expressed in different units, each variable was min-max rescaled to 0,1. Finally, atypicality is the Euclidean distance of the two scaled deviations, and the formula was fitted as follows:

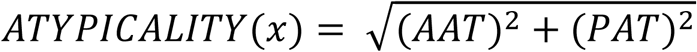

Where *x* is a given subject, AAT is rescaled amplitude atypicality, and PAT is rescaled acrophase atypicality. We compared the CSS with atypicality and other individual components metrics (e.g., amplitude atypicality and acrophase atypicality) in our framework. All the analyses were conducted in R Studio (R v4.3.1).

### 2.4 Use cases: CSS, Social Disadvantage, and Gut Microbiome

We provide two use cases on how the CSS can be used in both psychology/endocrinology studies involving classical statistical approaches or in more complex frameworks for multi-omics or metagenomic investigations.

We used hand-curated data from the Early Life Adversity Biological Embedding and Risk for Developmental Precursors of Mental Disorders (eLABE) study, a well-characterized sub-cohort of the March of Dimes research program. The eLABE cohort includes a total of 399 pregnant women who provided salivary cortisol samples during pregnancy underwent detailed psychosocial assessments, and demographic data collection across multiple stages of pregnancy [12,20]. As a validation step, we benchmarked the CSS against cortisol daily cycle state-of-the-art metrics by testing and comparing their associations with Social Disadvantage (SD)(as described above). To be consistent with the analyses, we included in the analysis only eLABE subjects having 6 salivary cortisol measures across a period of 24-hours (n_subjects_ = 196; n_samples_ = 1176) and implemented Spearman correlations between several cortisol-cycle-related variables (e.g., AUCg, MESOR, cosinor-estimated amplitude, JTK_CYCLE-estimated amplitude, atypicality, amplitude atypicality, acrophase atypicality) and SD.

Additionally, we provided an example of how cortisol daily cycle information can be used in gut microbiome studies by assessing the ability of the CSS to predict gut microbiome composition using a custom machine learning framework with the same parameters as in a previous eLABE microbiome study[21]. Broadly, we used a Random Forest regressor implemented with an 80/20 training and testing set split, repeated 100 times (10-fold, 10 times cross-validation) and trained on microbial species-level features only. Fecal sample collection and storage, data cleaning, biofiltering, preprocessing, and gut microbial species identification pipelines and protocols have been previously implemented and published in Warner, Rosa et al, 2023 [22].

## 3. Results

### 3.1 Minimum number of samples required for CSS estimation

We systematically evaluated all 63 possible combinations of the 6 daily cortisol time points from the 3006 samples to determine the minimum sampling protocol required to estimate the CSS accurately. We identified seven combinations that maintained high fidelity when compared to the full 6-sample CSS gold standard using stringent performance criteria (Pearson r > 0.9, MAE < 0.1) (**Figure 1A**, **Supplementary Table 1**). Analysis of time point importance revealed that the morning peak measurement at 06:00 and the evening measurement at 18:00 were the most critical ones, appearing in all seven of the high-performing combinations (**Figure 1B**). The mid-morning measurement at 10:00 was also highly important (6 occurrences), followed by the nadir at 02:00, afternoon at 14:00, and late evening at 22:00 (4 occurrences each). The combinations lacking the 06:00 time point sample, including the 5-sample combination (r > 0.69, MAE < 0.3), showed substantially reduced correlation with the full CSS (**Figure 1, Supplementary Table S1**), confirming that the morning cortisol peak is crucial for accurate CSS quantification. The minimum number of samples required to achieve acceptable performance (r > 0.9, MAE < 0.1) was 3 time points. The optimal 3-sample protocol consisted of measurements at 02:00, 06:00, and 18:00 (r = 0.952, MAE = 0.087), capturing the nocturnal nadir, morning peak, and evening decline. However, a 4-sample protocol using 06:00, 10:00, 14:00, and 18:00 achieved superior performance (r = 0.953, MAE = 0.023), providing more robust rhythm assessment while still reducing participant burden by 33% compared to the full 6 time point sampling protocol. Five-sample combinations achieved near-perfect agreement with the full CSS (r = 0.985, MAE < 0.093). These findings demonstrate that the CSS can be reliably estimated with as few as 3-4 daily measurements, substantially reducing participant burden while maintaining robust circadian rhythm quantification.

**Figure 1.**
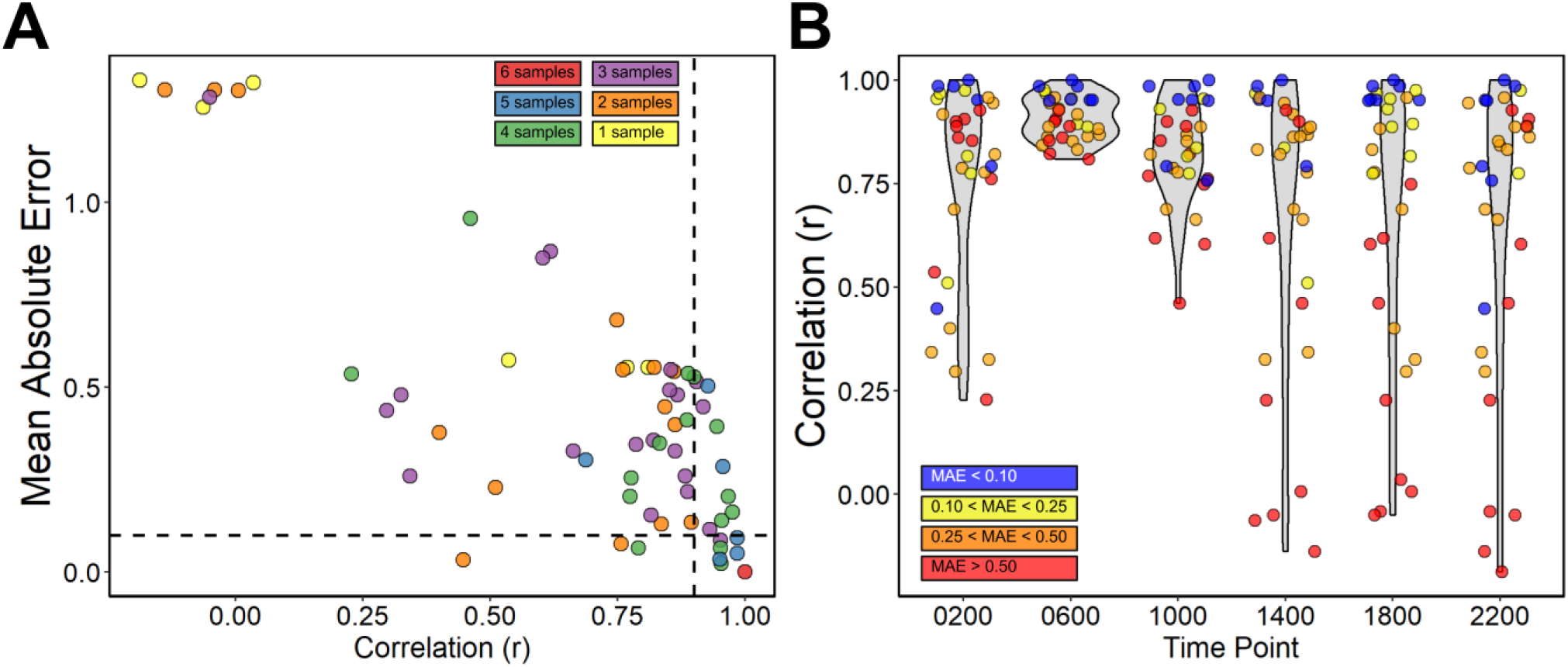
The minimum number of samples required for a good approximation (r > 0.9, MAE < 0.1) of the CSS is 3, but all the combinations have different performance. (**A**) The correlation coefficient r and MAE of the 63 possible combinations of CSS estimated from 3006 samples with 6-1 data points, and tested versus the 6-samples combination (red) after min-max rescaling of -1 to 1; (**B**) Violin plot of the correlation coefficient r and the MAE values of each specific time-point (e.g., in 7 different combinations, the timepoint 06:00 performed with r > 0.9, MAE < 0.1).

### 3.1 The CSS is associated with cortisol Amplitude and Acrophase

To validate and benchmark this novel metric, we compared the CSS against two widely used approaches: cosinor-regression and JTK_CYCLE. We evaluated CSS performance in recapitulating the rhythmic features defined by Cosinor and JTK_CYCLE including amplitude and acrophase (**Figure 2**). The CSS is a simple scalar metric, the Cosinor model is a parametric curve-fitting approach, while the JTK_CYCLE method is a non-parametric statistical test developed for rhythm detection. Because of the structural differences in the three different approaches, our validation efforts focused on assessing how well our simplified scalar metric approximates key rhythmic features captured by Cosinor and JTK_CYCLE.

**Figure 2.**
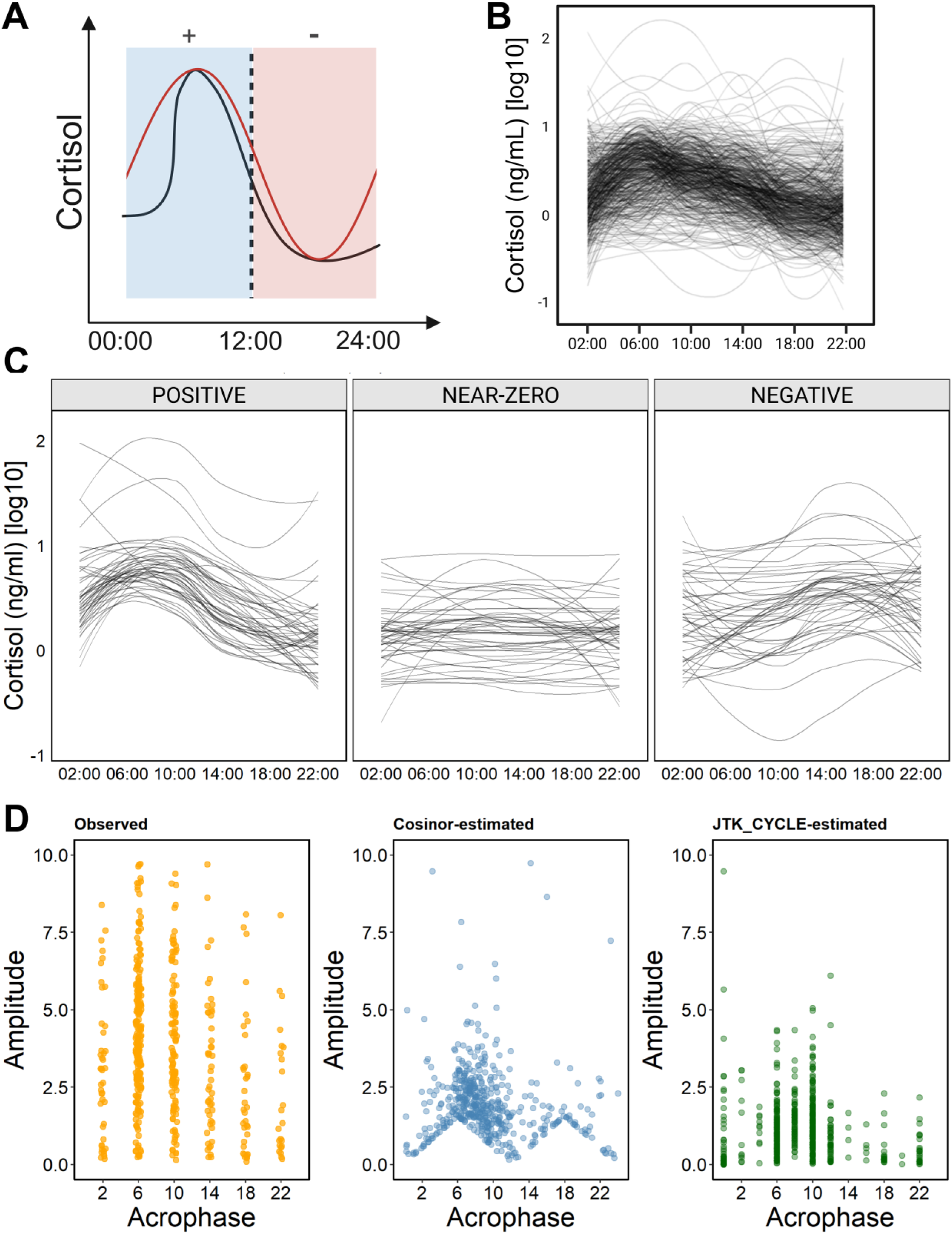
The relevance of CSS lies in its interpretability and usability: positive values reflect morning-aligned secretion, typical of a healthy cortisol daily cycle, negative values suggest delayed or inverted rhythms, while near-zero values indicate flattened or dysregulated patterns. (**A**) The sine function (red line) over a 24-hour period is employed as an approximation of the circadian rhythm (black line) of cortisol exposure and included in a dot product between the individual’s cortisol time series and a canonical 24-hour sine wave. Positive values are obtained if cortisol peaks together with the sine function (morning), negative values are obtained if cortisol peaks when the sine dips (evening). (**B**) LOESS curves (span=1) depicting the individual’s cortisol daily cycle of 501 subjects from the March of Dimes cohort. (**C**) LOESS curves (span=1) of cortisol daily cycle of the top (positive, n=50, range=[7.04,152.27]), bottom (negative, n=50, range=[- 62.72,-63.00]) and near-zero (n=50, range=[-0.24,0.23]) CSS values of 150 individuals from the March of Dimes cohort selected for demo purposes. (**D**) Comparison between observed (orange), Cosinor-estimated (blue) and JTK_CYCLE-estimated (green) Amplitude and Acrophase of 501 women from the March of Dimes cohort, during the second trimester of pregnancy.

The validation of CSS includes 3006 salivary cortisol samples from 501 subjects of the March of Dimes cohort. We first fitted linear mixed models and tested for associations between CSS and observed and estimated amplitude (via cosinor and JTK_CYCLE approaches). Regardless of the variable used, all the models consistently showed a strong association between the CSS and both observed and estimated amplitude (LM_obs_amp_: β=0.73, p < 0.0001; LM_cosinor_amp_ : β=2.08, p < 0.0001; LM_jtk_amp_: β=2.07, p < 0.0001; **Figure 3A, Supplementary Table S2**). Additionally, CSS successfully classified individuals as having low (Q1) or high (Q4) cortisol amplitude with mean AUC values of at least 0.88 across 3 methods (**Figure 3B**).

**Figure 3.**
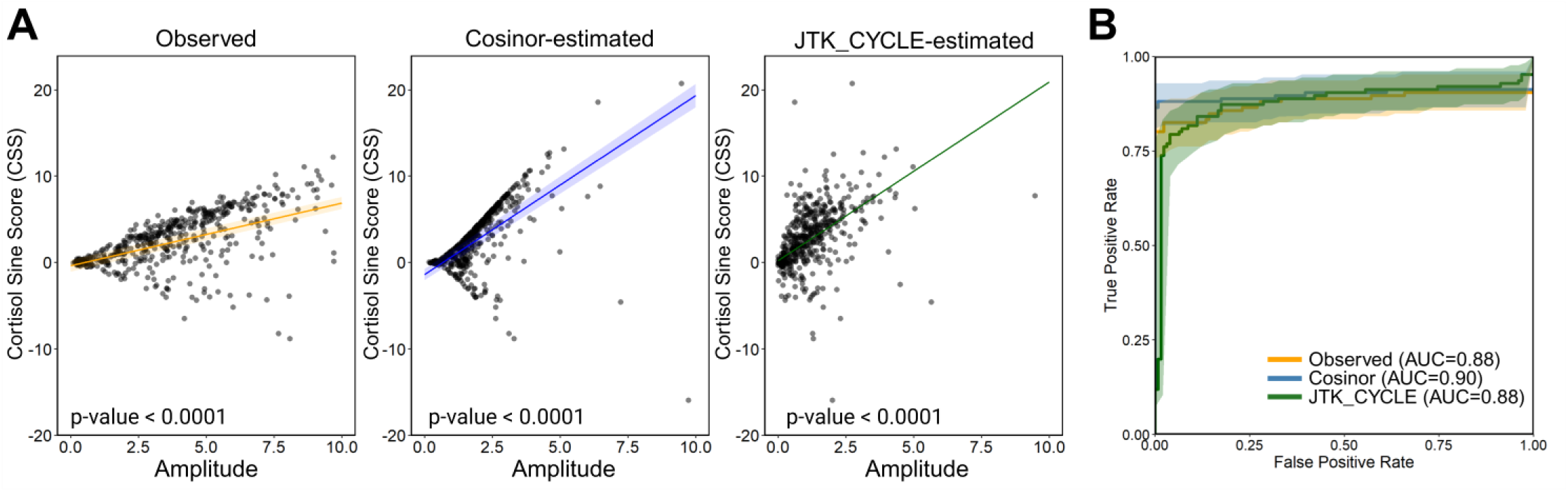
(**A**) Observed (orange), cosinor-estimated (blue) and JTK_CYCLE-estimated (green) cortisol Amplitude are positively associated with CSS (grey area = 95% CI); (**B**) ROC curves plotted according to true positive (recall) and false positive rates (precision) with corresponding AUC showing the discrimination between high (Q4) vs low (Q1) Amplitude (orange: Observed Amplitude AUC = 0.88; blue: Cosinor-estimated Amplitude AUC = 0.90; green: JTK-CYCLE-estimated Amplitude AUC = 0.88).

Second, we tested for associations between CSS and acrophase (observed and estimated) with two different statistical approaches: linear mixed models and general additive models. We fitted general additive models to account for the cyclical nature of the Acrophase time variable. Consistently, the results from both the LM and the GAM models showed a significant association between the CSS and the acrophase of cortisol exposure. Specifically, LMs showed that the CSS tertile categories had significantly different acrophases characterized by a shift in the cortisol peak hour of approximately 5 hours, from 07:00 to 13:00 (LM_obs_acrophase_: p < 0.0001; LM_cosinor_acrophase_ : p < 0.0001; LM_jtk_acrophase_ : p < 0.0001)(**Supplementary Table S2**). Similarly, the GAM analyses with cyclic cubic splines indicated that the CSS exhibits a clear sinusoidal pattern, with a significant acrophase effect observed across all models (GAM_obs_acrophase_: p<0.0001; GAM_cosinor_acrophase_: p<0.0001; GAM_jtk_acrophase_ : p<0.0001, **Supplementary Table S3**).

Overall, these results show that the CSS successfully captures components of both cortisol amplitude and phase (calculated using several models) and demonstrate its value to model cortisol cycle information condensed into one single metric applicable in wide range biomedical studies, such as endocrinology, psychology, and multi-omics studies.

### 3.3 Use cases: CSS, Social Disadvantage, and Gut Microbiome

The CSS is a simple scalar metric capturing alteration in the cortisol daily cycle making it applicable for use not only in classical statistical approaches but also in complex ML/AI frameworks more commonly used in multi-omics or metagenomics studies. Here, we provide two case studies, an example of how the CSS metric can be used in biomedical studies by using hand-curated data from the eLABE cohort (sub-cohort of the March of Dimes Prematurity Research Cohort Study). We benchmarked the CSS, and the CSS with the minimum sample size, against other cortisol cycle-related variables (**Figure 5** for complete list) with (a) Social Disadvantage (via Spearman correlations) as a crucial benchmark variable considering its well-established relationship with alteration in cortisol circadian rhythmicity and daily cycle; and (b) provided an example on how to use it in ML/AI frameworks (via Random Forest) for gut microbiome composition study for a demonstration of its use in multi-omics studies (**Figure 5**).

**Figure 4.**
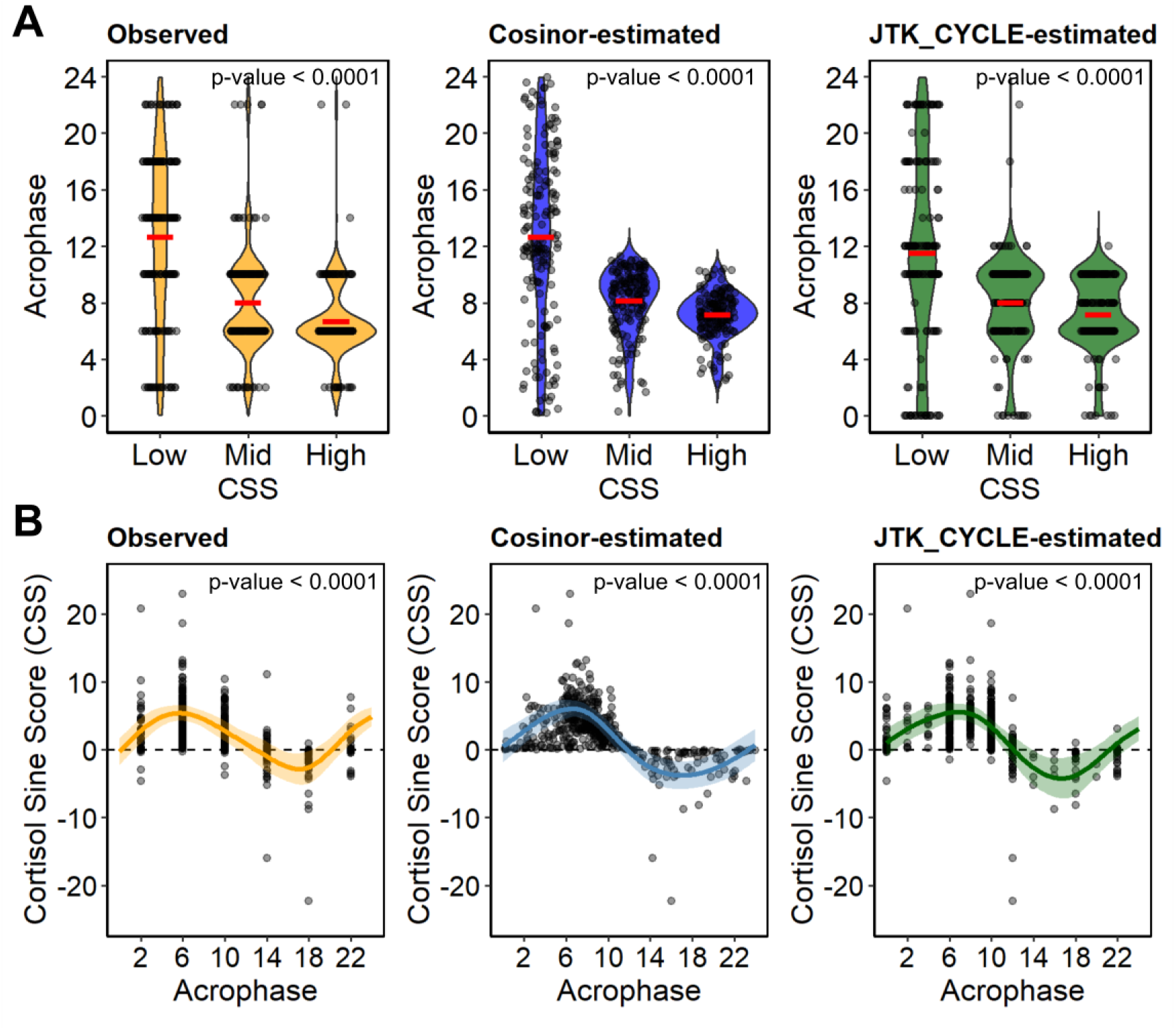
Observed (orange), cosinor-estimated (blue) and JTK_CYCLE-estimated (green) cortisol peak hour (Acrophase) is significantly associated with CSS. (**A**) Violin plots of CSS tertiles (low, mid high) plotted according with Acrophase show significant differences in peak hours (mean group value = red line). (**B**) Models line showing the predicted effect of the cortisol acrophase coded as a cyclic variable and explaining the CSS (colored areas = 95% CI).

**Figure 5.**
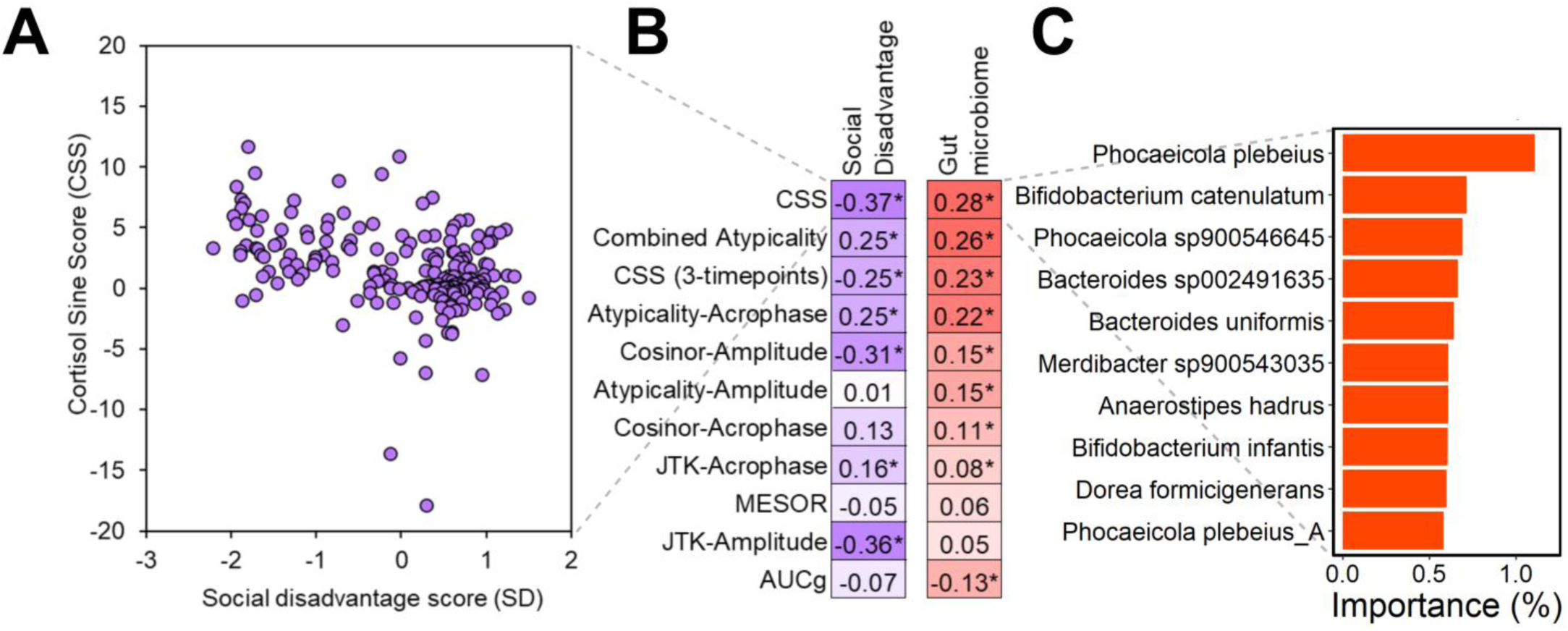
Cortisol Sine Score (CSS) use cases in psychology and multi-omics studies. (**A**) Cortisol Sine Score vs Social Disadvantage (SD) showing that higher SD is significantly (Spearman’s ρ=-0.37, Spearman’s p-value < 0.0001) associated with a lower CSS, which expresses a more flattened/blunted cortisol daily cycle. (**B**) Benchmarking of the Cortisol Sine Score (CSS) vs other cortisol daily cycle variables in a Spearman correlation (magenta, Social Disadvantage) and in a Random Forest (red, gut microbiome) framework shows the CSS as the best performing variable. * Indicate p-value < 0.05. (**C**) Top 10 gut microbial species associated with increasing CSS (ordered by mean feature importance).

The Spearman correlation values confirmed the proven link between Social Disadvantage and alterations of cortisol daily cycle, and the CSS resulted as the variable with the strongest associations in our benchmark framework (Spearman’s ρ=-0.37, Spearman’s p-value < 0.0001 (**Figure 5A**; Complete list of correlations vs other metrics in **Figure 5B**), followed by JTK and Cosinor estimated amplitude metrics (ρ=-0.37, ρ=-0.31). Interestingly, the 3-sample combination CSS (0200, 1000, 1800) performed better or equal to most of the state-of-the-art metrics (Spearman’s ρ=-0.25). In the second use case, when using a Random Forests regressor with 10- fold 10-times cross-validation approach trained on the gut microbiome species-level taxonomic profiles, the CSS performed as the best predictor of gut-microbial composition alterations (**Figure 5B**), followed by the Combined Atypicality, and by the CSS estimated using only 3 samples (0200, 1000, 1800). While two measures of amplitude (Cosinor and JTK) showed only slightly lower correlations with SD (-0.31 and -0.36), they both performed substantially worse in the gut microbiome comparison, showing the superiority of CSS across datasets. “Combined atypicality”, which was generated as an alternative but relatively simplistic single-value cortisol metric for phase and amplitude for comparison, was also outperformed by CSS in both tests and showed relatively comparable results in the gut microbiome testing. The top three among the most important gut microbial species associated with CSS were: *Phocaeicola plebeius* (mean importance = 1.1%), *Bifidobacterium catenulatum* (mean importance = 0.7%), and *Phocaeicola sp900546645* (mean importance = 0.7%) (**Figure 5C**).

Overall, these results confirmed that both the CSS and the reduced CSS (estimated with only 3 samples) have a strong value in expressing cortisol daily cycle disruption while collapsing information in a single scalar metric useful for classical statistical tests or ML/AI approaches complementary to existing approaches.

## 4. Discussion

The Cortisol Sine Score (CSS) represents a novel metric that characterizes diurnal cortisol patterns and offers a simple yet biologically meaningful scalar metric that addresses a critical gap in the field. The CSS collapses the temporal dynamics of cortisol exposure into a single interpretable value, providing clinicians and researchers with a metric for quick screening and capturing overall trends in HPA-axis functionality, while maintaining strong validity with well-established circadian rhythm analysis approaches.

Importantly, the CSS is designed to expand, but not replace, existing approaches like Cosinor regression and JTK_CYCLE. While these methods provide comprehensive multi-parameter characterization essential for detailed circadian rhythm analysis, the CSS enables rapid assessment of cortisol alignment and facilitates integration into statistical and ML/AI pipelines. Furthermore, its methodological simplicity requiring only a weighted sum calculation makes it easy to use without a need for specialized expertise in circadian analysis. The open-source R package *CortSineScore* facilitates an easy access to this tool.

The CSS is particularly valuable for initial screening, large-scale epidemiological studies, and scenarios where a quick assessment of overall cortisol rhythm integrity is needed before pursuing more detailed circadian analyses. Our validation demonstrates that the CSS successfully captures key features of the cortisol daily cycle. The strong associations with both observed and model-estimated Amplitude and Acrophase, combined with excellent discriminative ability (mean across three AUC = 0.89), confirm that this simplified metric retains essential biological information. Its directional nature provides immediate interpretability with positive values indicating morning-dominant cortisol exposure consistent with healthy circadian alignment, while negative values suggest evening-shifted patterns. Values approaching zero are produced by equal cortisol exposure across the day which translates into flattened or blunted rhythms, patterns consistently linked to chronic stress and detrimental health outcomes [3,8,18]. The ability to distinguish those patterns and their user-friendly accessibility is crucial for advancing our understanding of HPA-axis function across diverse populations and research contexts where sophisticated rhythm analysis may be impractical.

The CSS demonstrated great performance in capturing the well-established relationship between social disadvantage and cortisol dysregulation, outperforming other single or composite metrics in our benchmarking framework. Furthermore, its successful application in predicting gut microbiome composition illustrates its utility in complex metagenomics and multi-omics analyses where traditional multi-output methods performed worse and would be computationally prohibitive or conceptually inconvenient. This divergence likely reflects fundamental differences in what each metric captures. Cosinor and JTK_CYCLE amplitudes quantify the strength of rhythmic variation but are insensitive to its phase alignment or directionality. In contrast, the CSS integrates both amplitude and timing into a single scalar value that encodes the degree of morning versus evening dominance in cortisol exposure. Because microbial community structure is influenced by the timing and phase coherence of host circadian signals rather than amplitude alone, the CSS may serve as a more biologically relevant indicator of systemic circadian alignment. This finding highlights the advantage of CSS as a cross-domain biomarker that bridges endocrine rhythmicity with downstream physiological systems.

A crucial advantage of the CSS is its remarkable robustness to perform better or equally well using reduced sampling protocols. Our systematic evaluation of all possible time-point combinations revealed that the CSS can be reliably estimated with as few as 3-4 daily measurements while maintaining excellent agreement with the full 6-sample protocol (r > 0.90, MAE < 0.10). This finding has profound implications for study feasibility and participant burden reduction. The optimal 4-sample protocol (06:00, 10:00, 14:00, 18:00) achieves near-perfect performance (r = 0.953, MAE = 0.023) while excluding the burdensome nocturnal (02:00) and late evening (22:00) collections, representing a 33% reduction in sampling effort. Even a minimal 3-sample protocol capturing the nocturnal nadir, morning peak, and evening decline maintains acceptable accuracy (r = 0.952, MAE = 0.087). Interestingly, the CSS-based estimation with only 3 daily samples (0200, 0600, 1800) showed comparable power in detecting associations with social disadvantage and with gut microbiome composition, making it very useful in contexts where night data collection may be prohibitive or hard to implement. This flexibility is particularly valuable in pediatric populations, elderly individuals, and field studies where intensive sampling is impractical or poses an excessive burden. The non-negotiable importance of the 06:00 morning peak measurement identified in our analysis provides clear guidance for protocol optimization, ensuring researchers can confidently design abbreviated sampling schemes without compromising data quality. This scalability enhances the CSS translational potential, making circadian cortisol assessment feasible in resource-limited settings, remote monitoring applications, and large-scale cohort studies where intensive biospecimen collection would be prohibitive.

However, we acknowledge important limitations of the CSS. By design, the CSS trades information richness for simplicity and cannot capture complex rhythm features that full parametric or non-parametric analyses can detect. Additionally, while the CSS provides a useful summary of cortisol exposure across the day, it should not be used as the sole indicator of circadian disruption in contexts requiring detailed phase or amplitude characterization. Hypothetically, pathological endocrine conditions such as Addison’s disease (primary adrenal insufficiency) and Cushing’s syndrome (hypercortisolism) would both present with near-zero CSS values due to loss of diurnal variation despite representing opposite extremes of cortisol production. In Addison’s disease, the severely diminished cortisol output results in flat, low-amplitude profiles[23,24], while Cushing’s syndrome is characterized by persistently elevated cortisol with loss of the normal evening nadir[25]. This underscores an important consideration: the CSS specifically quantifies rhythm directionality and alignment rather than absolute cortisol magnitude. Therefore, in clinical or research contexts, where distinguishing hypocortisolism/adrenal insufficiency and hypercortisolism is critical, the CSS should be interpreted alongside measures of total cortisol output such as AUCg or mean cortisol levels. This complementary approach enables differentiation between healthy circadian alignment (high positive CSS), chronic stress-related flattening (near-zero CSS), adrenal insufficiency (near-zero with low cortisol), and hypercortisolism (near-zero/negative CSS with elevated cortisol). Furthermore, CSS absolute value is not an absolute diagnosis indicator. As such, CSS values cannot be interpreted against an absolute “normal” threshold across individuals or studies. Instead, the CSS is best used for relative comparisons within a population or same-subject variation across time, where higher positive values indicate stronger morning dominance and more intact rhythmicity. Future research should prioritize validating the CSS across diverse populations and physiological states, including shift workers, individuals with jet lag, and those with documented circadian rhythm disorders. Exploring optimal sampling schedules for CSS calculation and investigating whether modified weightings could better capture specific types of dysregulations and enhance its utility.

Finally, the potential for real-time CSS monitoring using wearable biosensors represents an exciting translational opportunity for R&D departments and entrepreneurs. The mathematical simplicity of the CSS makes it ideally suited for implementation in resource-constrained wearable devices, unlike complex curve-fitting algorithms that demand substantial computational power. As continuous cortisol biosensors advance from prototype to market (including emerging sweat-based and microneedle technologies), the CSS could enable real-time feedback about HPA-axis alignment without requiring cloud processing or sophisticated onboard analytics.

The single-value output of CSS makes it particularly attractive for consumer-facing applications where complex rhythm parameters would overwhelm users, while its directional nature (positive/negative/near-zero) provides intuitive feedback that could guide behavioral interventions. For the digital health industry, the CSS offers a standardizable metric simple enough for mass adoption yet scientifically robust enough for clinical decision support.

In conclusion, the CSS bridges the gap between detailed circadian biology and large-scale data science applications by offering a practical, validated solution for integrating cortisol daily cycle information into modern analytical frameworks. While it does not replace comprehensive rhythm analysis, it serves as a complementary tool that accelerates discovery in stress physiology research, facilitates clinical screening for HPA-axis dysfunction, and enables new investigations on the cortisol daily cycle and complex health outcomes. Together, our work introduces and validates the CSS as a compact, interpretable, and practical alternative for modelling cortisol daily cycle patterns in large-scale studies and applied contexts where a single scalar input is preferred or required.

## Declaration of generative AI and AI-assisted technologies in the manuscript preparation process

During the preparation of this manuscript, the authors used Claude Opus 4.1 to improve readability and assist with coding and literature searches. After using Claude Opus 4.1, the authors reviewed and edited the content as needed and take full responsibility for the content of the published article.

## CRediT author statement

**Simone Anzà**: Conceptualization, Methodology, Software, Validation, Formal Analyses, Investigation, Visualization, Writing Original Draft, Writing Review & Editing; **Bruce A. Rosa**: Conceptualization, Methodology, Writing Original Draft, Writing Review & Editing, Supervision; **Max P. Herzberg**: Data Curation, Conceptualization, Writing Review & Editing; **Giljae Lee**: Writing Review & Editing; **Erik D. Herzog**: Conceptualization, Writing Review & Editing; **Peinan Zhao**: Conceptualization, Writing Review & Editing; **Sarah K. England**: Conceptualization, Writing Review & Editing; **Malick Ndao**: Data Curation; **John Martin**: Data Curation; **Christopher D. Smyser**: Writing Review & Editing, Funding acquisition, Project administration; **Cynthia E. Rogers**: Writing Review & Editing; **Deanna M. Barch**: Writing Review & Editing; **Caroline Hoyniak**: Writing Review & Editing; **Ronald McCarthy**: Data Curation; **Joan Luby**: Supervision, Writing Review & Editing, Funding acquisition, Project administration; **Barbara B. Warner**: Supervision, Writing Review & Editing, Funding acquisition, Project administration; **Makedonka Mitreva**: Supervision, Writing Review & Editing, Funding acquisition, Project administration.

## Data Availability Statement

The dataset includes sensitive and confidential patient information (e.g., salivary cortisol measurements, sociodemographic data, and clinical information) collected from pregnant women enrolled in the March of Dimes Prematurity Research Cohort Study. Data sharing would violate participant privacy protections and institutional review board restrictions. Researchers interested in accessing these data may contact the corresponding authors to discuss potential collaboration opportunities under appropriate data use agreements and institutional review board approvals.

## SUPPLEMENTARY INFO

**Supplementary Table S1.** Results from the correlations between each specific combination of timepoints and the 6-sample combination. MAE = mean absolute error.

| Combination<br>(n samples) | Timepoint included | Pearson's correlation<br>(r) | MAE |
| --- | --- | --- | --- |
| 1 | 0200 | 0.536565422 | 0.573086373 |
| 1 | 0600 | 0.809460526 | 0.553571563 |
| 1 | 1000 | 0.768651369 | 0.554060325 |
| 1 | 1400 | -0.063358667 | 1.257063953 |
| 1 | 1800 | 0.034950836 | 1.32371417 |
| 1 | 2200 | -0.188097657 | 1.331264303 |
| 2 | 0200,0600 | 0.860194476 | 0.542065847 |
| 2 | 0200,1000 | 0.760492034 | 0.546757199 |
| 2 | 0200,1400 | 0.510152632 | 0.228151499 |
| 2 | 0200,1800 | 0.399810521 | 0.378149354 |
| 2 | 0200,2200 | 0.447642134 | 0.033804347 |
| 2 | 0600,1000 | 0.821447787 | 0.553948766 |
| 2 | 0600,1400 | 0.862619171 | 0.399461514 |
| 2 | 0600,1800 | 0.894082072 | 0.135101653 |
| 2 | 0600,2200 | 0.842165391 | 0.447755969 |
| 2 | 1000,1400 | 0.835796392 | 0.12941389 |
| 2 | 1000,1800 | 0.748611619 | 0.682497688 |
| 2 | 1000,2200 | 0.757368998 | 0.076918838 |
| 2 | 1400,1800 | 0.005386162 | 1.302996297 |
| 2 | 1400,2200 | -0.138560752 | 1.304541748 |
| 2 | 1800,2200 | -0.041848108 | 1.304102566 |
| 3 | 0200,0600,1000 | 0.853956403 | 0.546932913 |
| 3 | 0200,0600,1400 | 0.917662575 | 0.447642622 |
| 3 | 0200,0600,1800 | 0.952037351 | 0.087050498 |
| 3 | 0200,0600,2200 | 0.90510919 | 0.51585419 |
| 3 | 0200,1000,1400 | 0.820438056 | 0.356008284 |
| 3 | 0200,1000,1800 | 0.816190851 | 0.154166575 |
| 3 | 0200,1000,2200 | 0.786846265 | 0.345988269 |
| 3 | 0200,1400,1800 | 0.324686174 | 0.478886356 |
| 3 | 0200,1400,2200 | 0.342960603 | 0.259707074 |
| 3 | 0200,1800,2200 | 0.296263779 | 0.437989378 |
| 3 | 0600,1000,1400 | 0.867901039 | 0.480065155 |
| 3 | 0600,1000,1800 | 0.930423473 | 0.116306644 |
| 3 | 0600,1000,2200 | 0.85191503 | 0.492689146 |
| 3 | 0600,1400,1800 | 0.882793389 | 0.259549125 |
| 3 | 0600,1400,2200 | 0.862919735 | 0.327316315 |
| 3 | 0600,1800,2200 | 0.886933343 | 0.217096132 |
| 3 | 1000,1400,1800 | 0.618862925 | 0.868486914 |
| 3 | 1000,1400,2200 | 0.663076868 | 0.32801393 |
| 3 | 1000,1800,2200 | 0.603101697 | 0.850336519 |
| 3 | 1400,1800,2200 | -0.050572658 | 1.285855016 |
| 4 | 0200,0600,1000,1400 | 0.899996027 | 0.528641691 |
| 4 | 0200,0600,1000,1800 | 0.954438635 | 0.140057294 |
| 4 | 0200,0600,1000,2200 | 0.888397094 | 0.537807085 |
| 4 | 0200,0600,1400,1800 | 0.967461151 | 0.204670994 |
| 4 | 0200,0600,1400,2200 | 0.944863704 | 0.393507175 |
| 4 | 0200,0600,1800,2200 | 0.975338927 | 0.161740227 |
| 4 | 0200,1000,1400,1800 | 0.777207359 | 0.254232357 |
| 4 | 0200,1000,1400,2200 | 0.790992729 | 0.065658114 |
| 4 | 0200,1000,1800,2200 | 0.774749615 | 0.20443043 |
| 4 | 0200,1400,1800,2200 | 0.227642182 | 0.536173494 |
| 4 | 0600,1000,1400,1800 | 0.953320649 | 0.023171907 |
| 4 | 0600,1000,1400,2200 | 0.8865925 | 0.411623336 |
| 4 | 0600,1000,1800,2200 | 0.951883653 | 0.065788695 |
| 4 | 0600,1400,1800,2200 | 0.831656526 | 0.348439976 |
| 4 | 1000,1400,1800,2200 | 0.461100326 | 0.956874602 |
| 5 | 0200,0600,1000,1400,1800 | 0.985083044 | 0.050913345 |
| 5 | 0200,0600,1000,1400,2200 | 0.927430128 | 0.50335645 |
| 5 | 0200,0600,1000,1800,2200 | 0.985296555 | 0.092765092 |
| 5 | 0200,0600,1400,1800,2200 | 0.956852644 | 0.285439214 |
| 5 | 0200,1000,1400,1800,2200 | 0.687266984 | 0.302940158 |
| 5 | 0600,1000,1400,1800,2200 | 0.950896425 | 0.034327871 |
| 6 | 0200,0600,1000,1400,1800,2200 | 1 | 0 |

**Supplementary Table S2.** Linear Mixed Models and variables used to validate the CSS against state-of-the-art approaches.

| Model | Variable | Estimate | Std. Error | Z-value | CI (2.5%;97.5%) | p-value |
| --- | --- | --- | --- | --- | --- | --- |
| Observed Amplitude (R<sup>2</sup>=0.45) | Intercept | -0.39 | 0.36 | -1.09 | -1.09;0.31 | - |
|  | CSS | 0.73 | 0.04 | 20.16 | 0.66;0.80 | **<0.001** |
| Cosinor-estimated Amplitude (R<sup>2</sup>=0.56) | Intercept | -1.41 | 0.33 | -4.29 | -2.06;-0.77 | - |
|  | CSS | 2.08 | 0.08 | 25.34 | 1.92;2.24 | **<0.001** |
| JTK_CYCLE-estimated Amplitude (R<sup>2</sup>=0.62) | Intercept | 0.22 | 0.28 | 0.81 | -0.32;0.76 | - |
|  | CSS | 2.07 | 0.07 | 28.74 | 1.93;2.21 | **<0.001** |
| Observed Acrophase (R<sup>2</sup>=0.26) | Intercept | 12.61 | 0.34 | 37.41 | 11.95;13.27 | - |
|  | CSS_Mid* | -4.62 | 0.48 | -9.7 | 5.56;-3.69 | **<0.001** |
|  | CSS_High* | -5.96 | 0.48 | -12.51 | -6.90;-5.03 | **<0.001** |
| Cosinor-estimated Acrophase (R<sup>2</sup>=0.27) | Intercept | 12.64 | 0.3 | 41.53 | 12.04;13.23 | - |
|  | CSS_Mid* | -4.51 | 0.43 | -10.49 | -5.36;-3.67 | **<0.001** |
|  | CSS_High* | -5.5 | 0.43 | -12.79 | -6.35;-4.66 | **<0.001** |
| JTK_CYCLE-estimated Acrophase (R<sup>2</sup>=0.15) | Intercept | 11.47 | 0.35 | 32.59 | 10.78;12.16 | - |
|  | CSS_Mid* | -3.51 | 0.49 | -7.05 | -4.48;-2.53 | **<0.001** |
|  | CSS_High* | -4.34 | 0.49 | -8.71 | -5.31;-3.36 | **<0.001** |

\* = low CSS tertile as reference category. Significant P values are in bold

**Supplementary Table S3.** General Additive Models and variables used to validate the CSS against state-of-the-art approaches.

| Model | Smooth | edf | df | F-value | p-value |
| --- | --- | --- | --- | --- | --- |
| Observed Acrophase<br>(R<sup>2</sup>adj=0.08) | CSS | 2.99 | 4 | 10.8 | <0.001 |
| Cosinor-estimated<br>Acrophase<br>(R<sup>2</sup>adj=0.12) | CSS | 2.82 | 4 | 16.4 | <0.001 |
| JTK_CYCLE-estimated Acrophase<br>(R<sup>2</sup>adj=0.08) | CSS | 2.94 | 4 | 10.4 | <0.001 |

